# Validation of sleep-based actigraphy machine learning models for prediction of preterm birth

**DOI:** 10.1101/2025.03.05.25323418

**Authors:** Benjamin C. Warner, Peinan Zhao, Erik D. Herzog, Antonina I. Frolova, Sarah K. England, Chenyang Lu

## Abstract

Disruptive sleep is a well-established predictor of preterm birth. However, the exact relationship between sleep behavior and preterm birth outcomes remains unknown, in part because prior work has relied on self-reported sleep data. With the advent of smartwatches, it is possible to obtain more reliable and accurate sleep data, which can be utilized to evaluate the impact of specific sleep behaviors in concert with machine learning. We evaluate motion actigraphy data collected from a cohort of participants undergoing pregnancy, and train several machine learning models based on aggregate features engineered from this data. We then evaluate the relative impact from each of these actigraphy features, as well as features derived from questionnaires collected from participants. Our findings suggest that actigraphy data can predict preterm birth outcomes with a degree of effectiveness, and that variability in sleep patterns is a relatively fair predictor of preterm birth.

## 1 Introduction

Preterm birth (PTB), which is generally defined as delivery before 37 weeks of gestation, is the single largest cause of death in children under the age of 5 [1] with approximately 1 million deaths occuring per year [2]. While some etiologies of PTB have been identified, many remain unknown. Previous literature has shown that disruptive maternal sleep patterns have been associated with PTB outcomes [3–6].

One major limitation with previous studies is the reliance on self-reported sleep patterns, which is limited by a patient’s ability to recall their sleep patterns accurately and consistently [7]. Wearable devices can alleviate this problem as they provide a more reliable and detailed stream of data [8, 9]. Previous literature has found that wearable sensor data can be used to make predictions regarding both physical and mental health issues, ranging from pancreatic complications [10] to depression [11].

Using data collected from wearables, we evaluate predictions of binary PTB outcomes with patients from a cohort study conducted at Washington University in St. Louis/BJC HealthCare [12]. Participants from this cohort study were given actigraphy watches to wear for two weeks over the course of each trimester, capturing high-resolution sleep data. The collected actigraphy data are then transformed into interpretable quantitative features and used as input for several shallow machine learning (ML) models. These models are then evaluated to assess the relative impact of these features, offering several clinical insights into the relative importance of individual sleep and non-sleep behaviors, as well as insights for more complex ML models.

Previous work with this dataset has attempted to evaluate regression models of unengineered time-series data to predict the entire spectrum of gestational age (GA) directly from individual actigraphy samples [13], which is intrinsically different in both objective and approach from predicting binary-outcome PTB from statistics across a pregnancy. The authors noted that measured mean absolute error between actual and predicted GA was higher overall in PTB patients, but did not evaluate any classifier performance with respect to binary-outcome PTB. Moreover, the models presented in [13] are limited in their explanability as a result of both learning non-linear representations and at attempting to predict GA at a sample level. In addition, previous work has also examined direct correlations between engineered actigraphy features and PTB, evaluating the risk associated with each individual feature [5, 6].

This paper evaluates the performance of binary-outcome classification of PTB from engineered actigraphy features and selected patient history features. The models presented here are computationally simpler and interpretable, which offer engineering and clinical insights about potential approaches for more complicated models. Overall, we validate the usage of sleep measures derived from actigraphy data in ML models for the prediction of binary-outcome PTB. From these models, relative comparisons of the impact of actigraphy and patient history features on predictions are examined. We finally offer interpretations of each of the tested models, and guidance for future works.

## 2 Results

Among the 1523 patients who participated in the cohort study, we analyze the 665 patients who had actigraphy data in at least the first or second trimester of their pregnancy and had a recorded delivery date. The average patient had 39.1 (*±*32.2) day-level samples throughout the duration of their pregnancy, with the first trimester having 15.7 (*±*10.4) samples on average, the second trimester having an average of 24.0 (*±*18.9) samples, and the third trimester having an average of 17.0 (*±*10.3) samples. The overall distribution of samples collected from all patients can be seen in Figure 1. Of these patients, the mean age was 29.2 (*±*5.29) years, and the majority (55.34%) of the patients were multiparous. A minority of patients (14.18%) experienced a PTB outcome. Full details about the demographics of the patients used in this dataset can be found in Section 1 in the Supplementary Materials.

**Figure 1:**
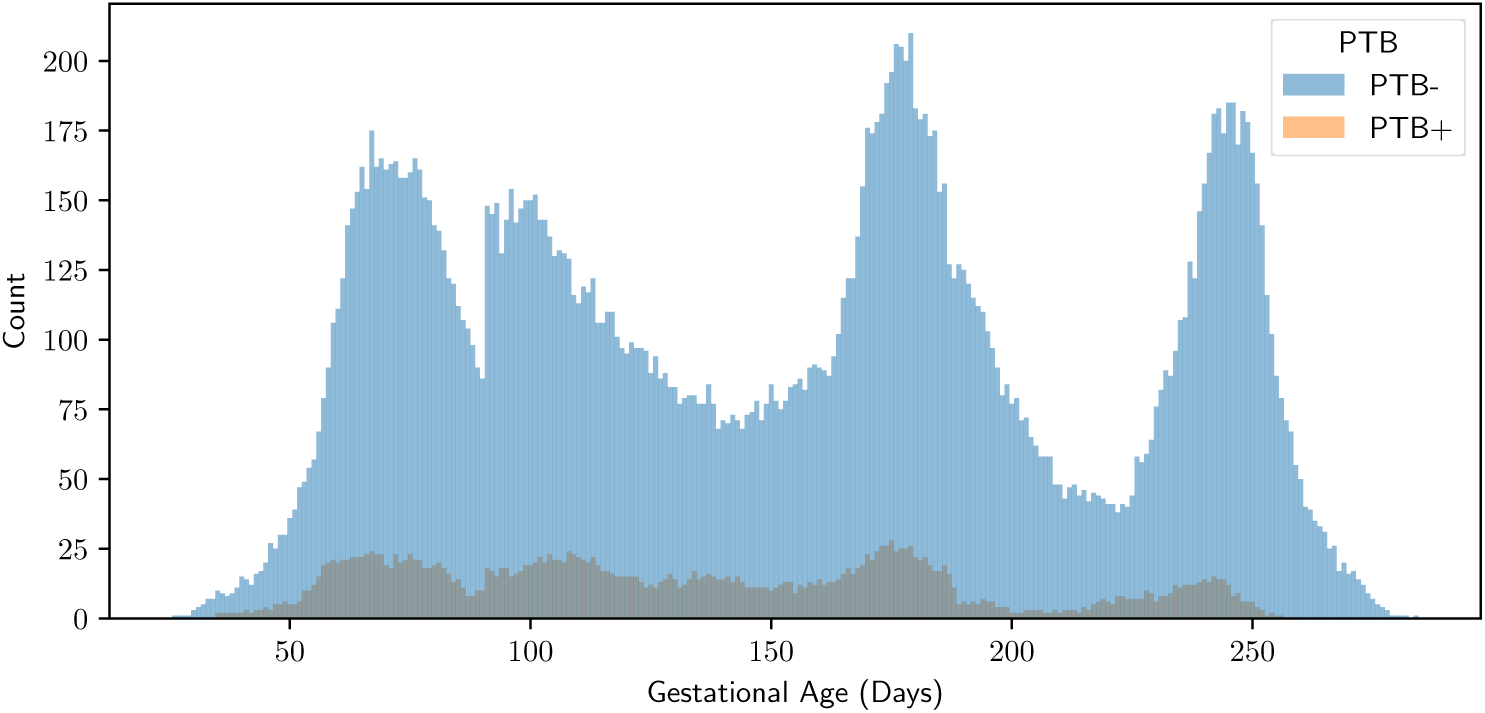
Histogram of the collected actigraphy samples. Results are stratified by whether the patient experienced a positive or negative PTB outcome.

We compare the performance of models trained on the two primary sources of data, the engineered actigraphy features and case report form responses collected at each visit, in Table 1 and Figures 2 to 4. We find that, using actigraphy features and case report form survey data, it is possible to make reasonable predictions about binary-outcome PTB. As seen in Table 1 and Figure 3, actigraphy features appear to underperform features from case report forms at predicting PTB when comparing the best models for each configuration. The combined performance is better than either source of data individually.

**Figure 2:**
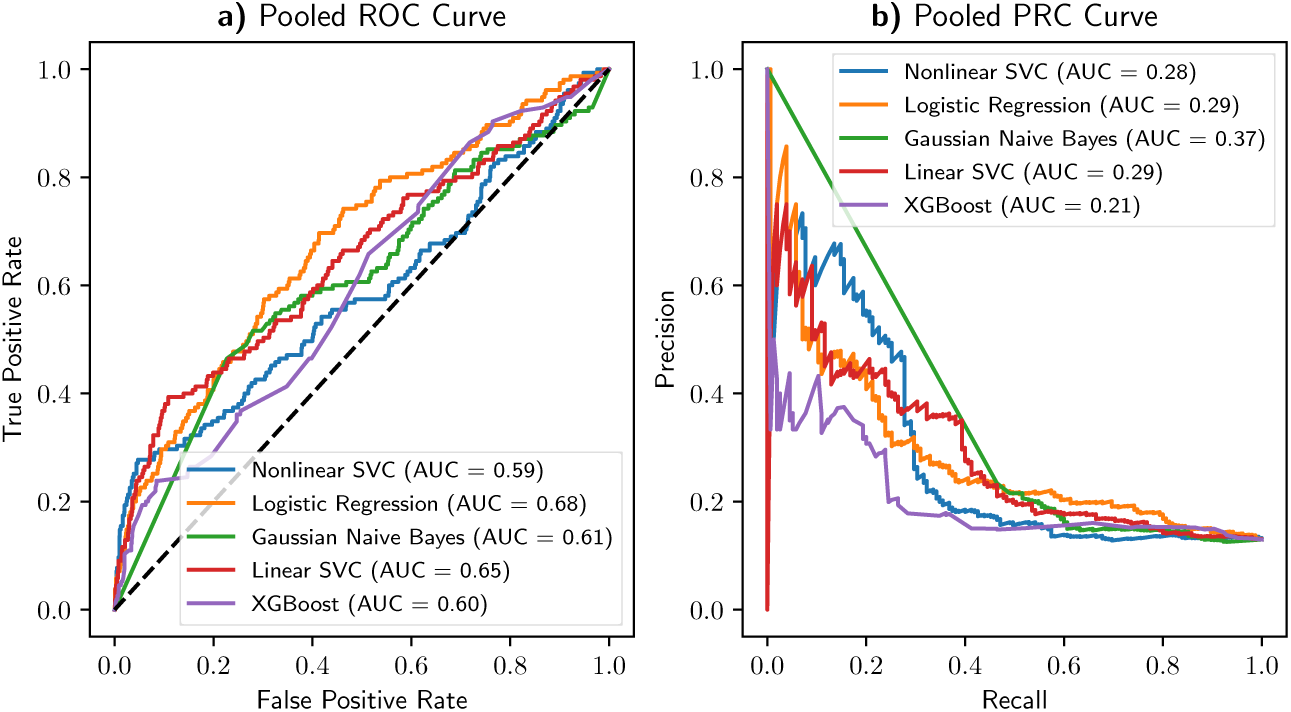
Reciever-operator and precision-recall curves for models using all features. Selected a) ROC curves and b) PRC curves for the median performing models using all data sources.

**Figure 3:**
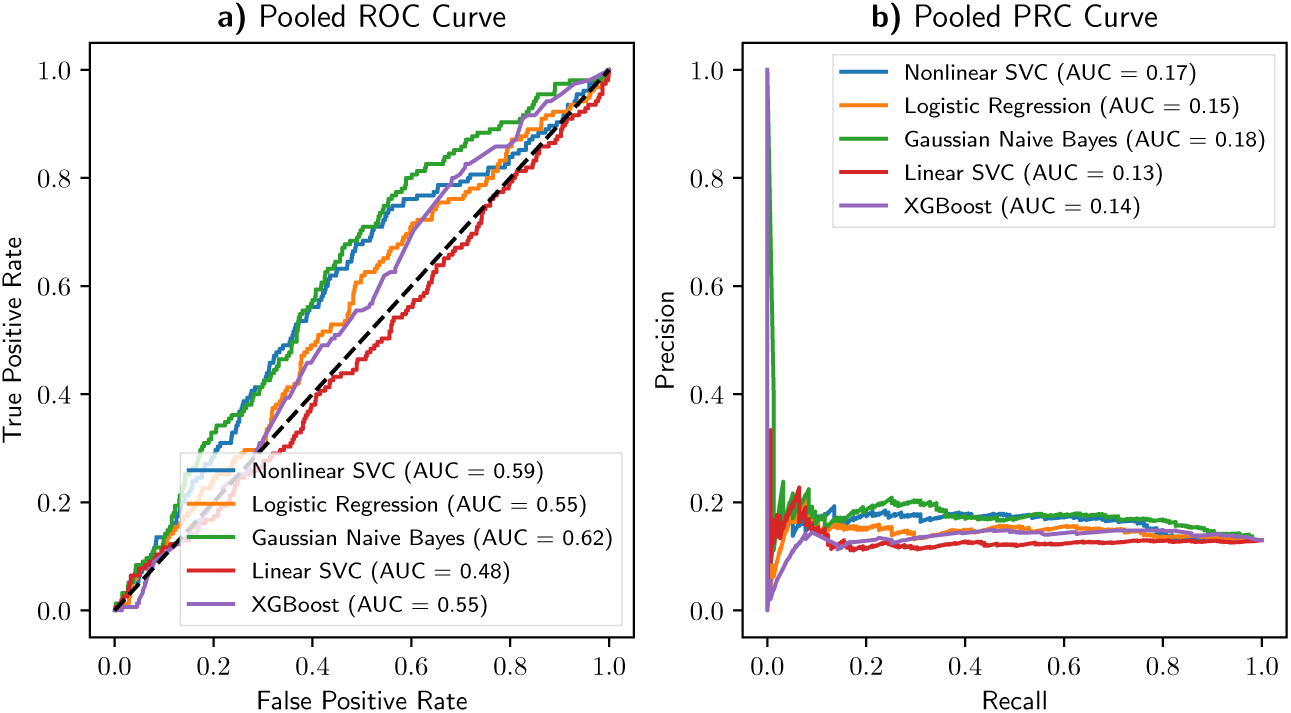
Reciever-operator and precision-recall curves for models using actigraphy data only. Selected a) ROC curves and b) PRC curves for the median performing models using only the actigraphy data.

**Figure 4:**
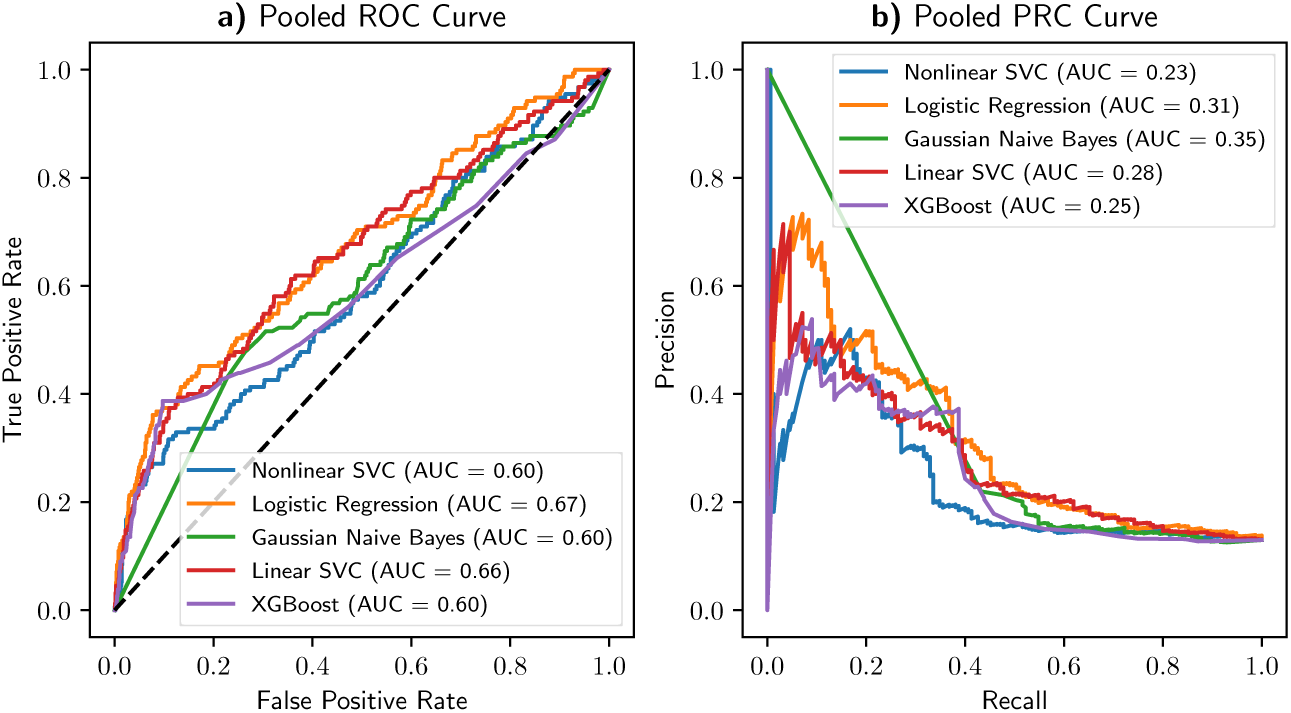
Reciever-operator and precision-recall curves for models using case report form data only. Selected a) ROC curves and b) PRC curves for the median performing models using only the case report form data.

**Table 1:**
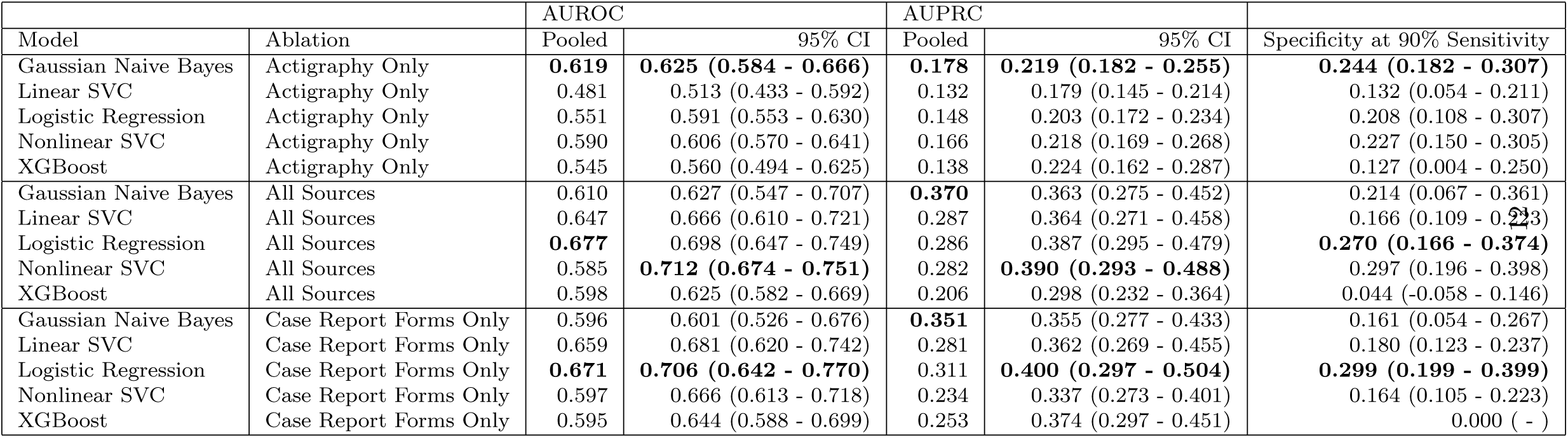
Comparison of models for all patients. AUROC, AUPRC, and specificity at 90% sensitivity highlighted for each of the trained models, grouped by which sources of data were included. Averages for AUROC and AUPRC are obtained by pooling classifier results together, and 95% confidence intervals are obtained by averaging all folds.

### 2.1 Gestational age and Model Performance

Figure 5 shows the performance of each model as samples up to a specified GA are included. As seen, the performance of the models does not change consistently as the GA upper-bound is increased, although it does increase noticeably in performance as the full GA spectrum is enabled.

**Figure 5:**
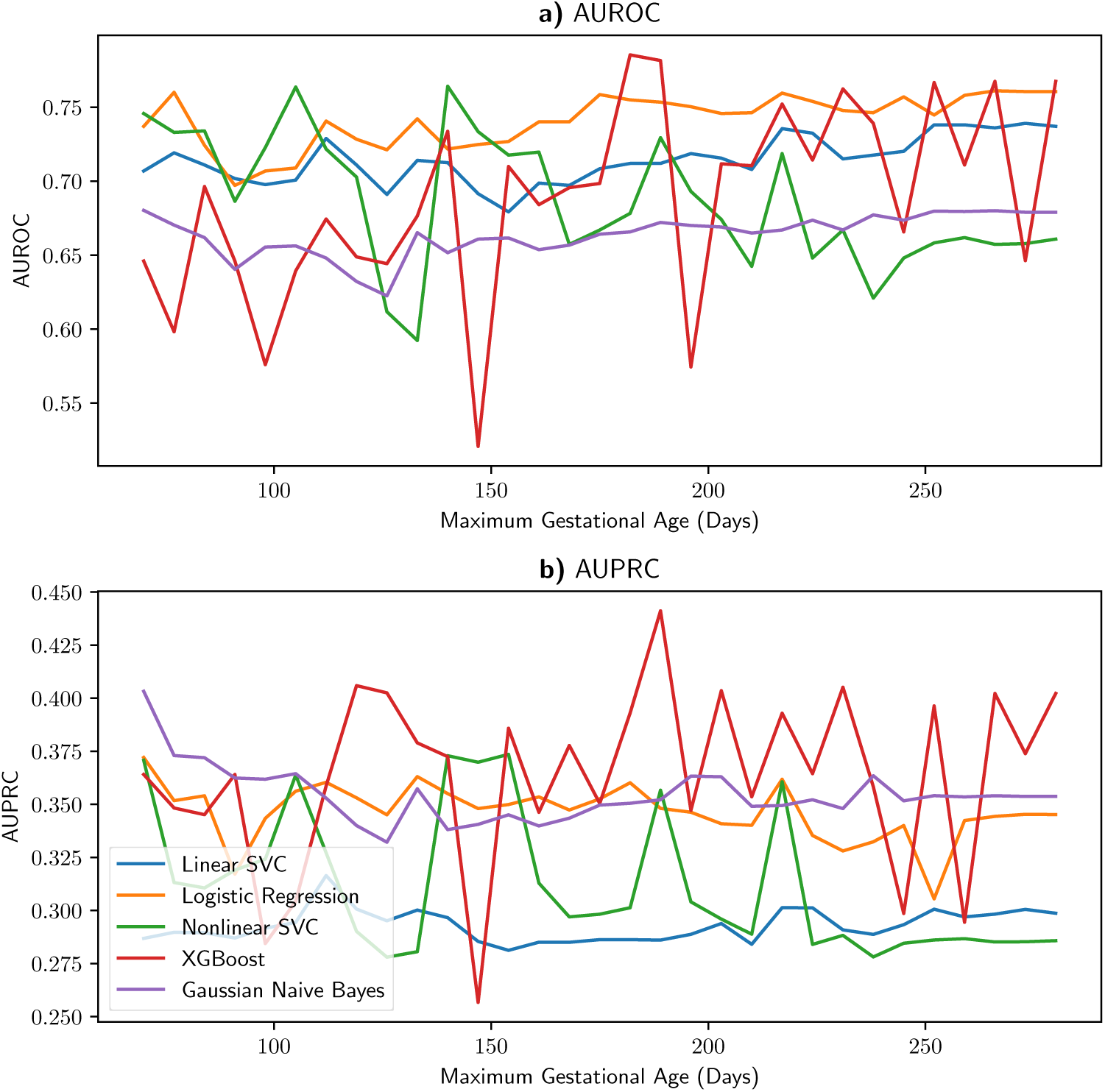
Reciever-operator and precision-recall curves for data up to a given GA. Selected a) ROC curves and b) PRC curves using with all features calculated with features up to a maximum GA using one random seed.

This lack of consistent performance change likely occurs for several reasons. First, the distribution of study participants who have data up to a given GA is variable, and for those that do have data up to a specified GA, the duration and lengths are also variable. In addition, the aggregation used for all actigraphy features, mean and standard deviation, does not change linearly as the amount of data increases. This variability in area under the receiver-operator curve (AUROC) and area under the precision-recall curve (AUPRC) appears to weakly correspond to the sample trends seen in Figure 1, which is roughly centered around the boundaries in each trimester.

### 2.2 Feature Explanations

To assess the importance of each feature in each model, we evaluate the features with SHapley Additive exPlanations (SHAP) scores [14], which provide relative estimates of how the output of a model will change as the input features change. Figure 6 shows the feature explanations for the best performing model with all features.

**Figure 6:**
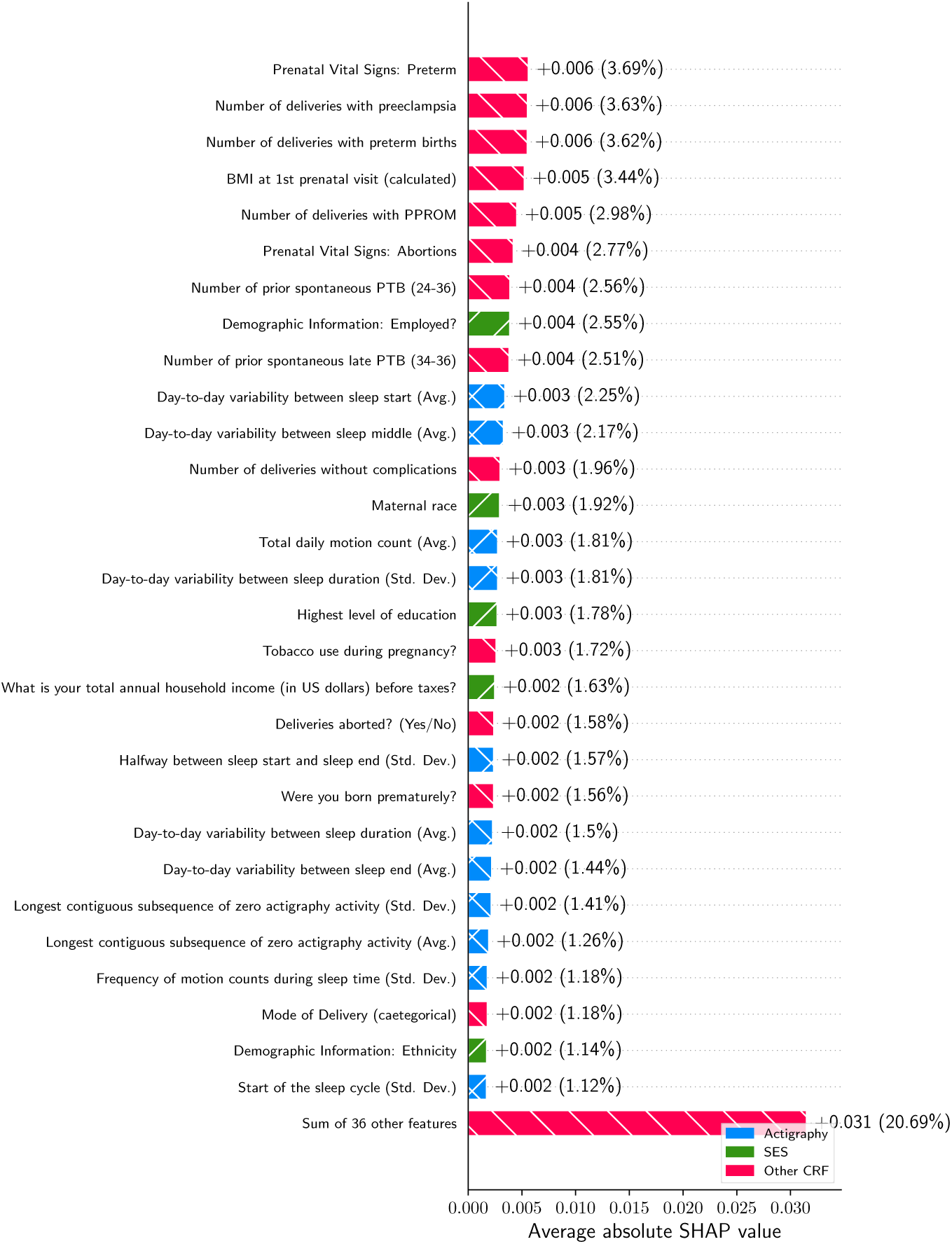
SHAP analysis of logistic regression with all features. Features indicative of socioeconomic status are highlighted green, and other patient history variables are highlighted red.

When all features are used, we find that the features that affect the output of the model the most are related to the number of complications that occurred during previous births. This is consistent with the literature, which finds that past PTB is a strong predictor of future PTB outcomes [15, 16]. Features relating to socioeconomic status, highlighted in green, also rank highly, which is consistent with the literature [17, 18].

Actigraphy features were impactful to a lesser degree, with the highest ranked feature being the average day-to-day variability between sleep start. Other similarly ranked features following this included sleep start time, the variance of the start of the sleep cycle, and day-to-day variability in the duration of the sleep cycle, *etc.* Overall, actigraphy features relating to variability in sleep patterns appeared to rank higher than those derived from averages across a patient’s pregnancy.

When we evaluate the best performing actigraphy-only model, shown in Figure 7, we find a similar ordering of relevant features, with features reflecting variance between daily actigraphy measurements appearing towards the top. Some of this difference in ordering can be attributed to the issue of dimensionality, as the number of examples is smaller, although the limited sample size prevents us from making any conclusive orderings.

**Figure 7:**
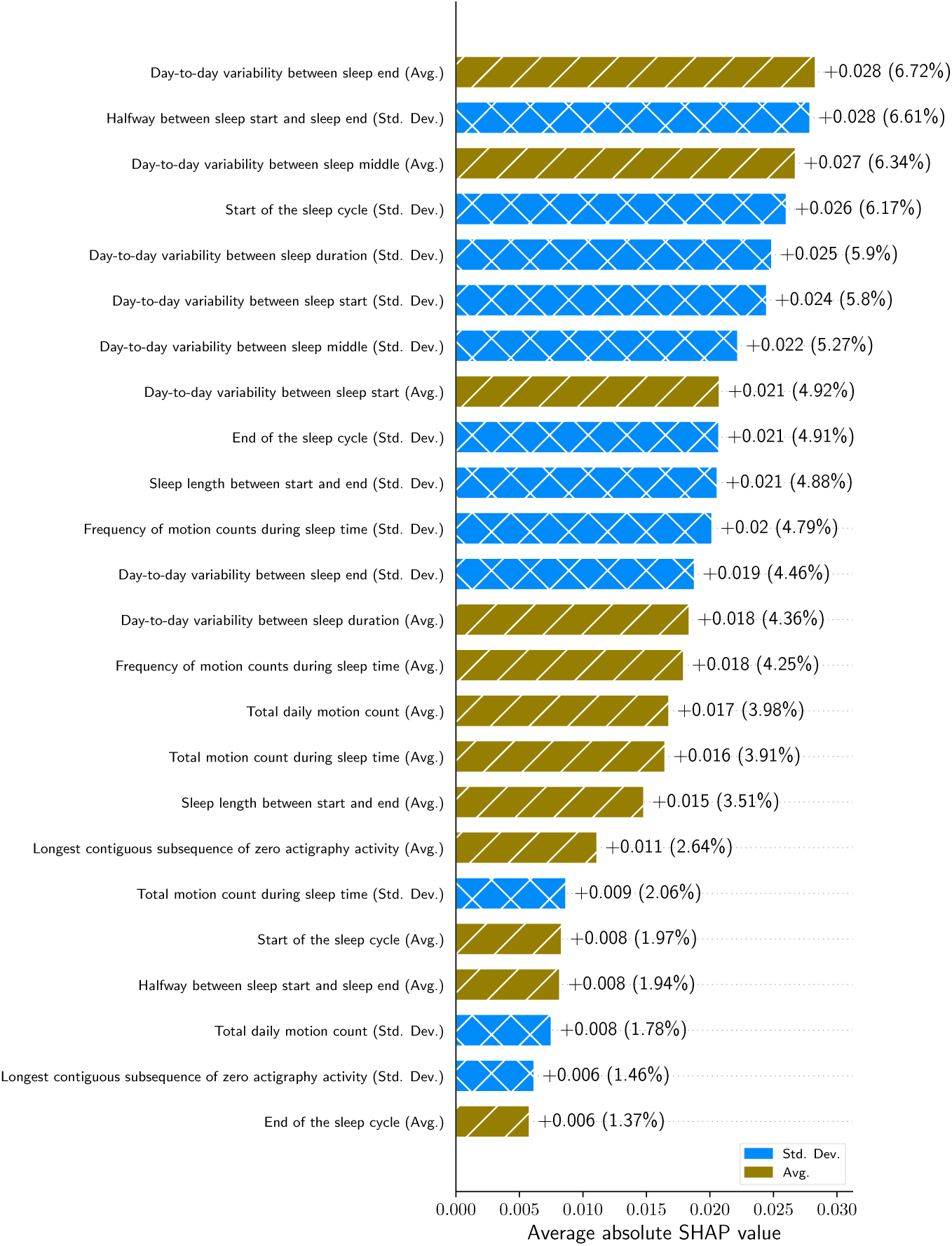
SHAP analysis of Gaussian NB with actigraphy features only. Features aggregating average patient behavior are highlighted in gold, and features aggregating the standard deviation of patient behaviors are highlighted in blue.

### 2.3 First-Time/Nulliparous Pregnancies

Prior PTB complications are a strong predictor of future PTB complications, but such foresight does not exist in the case of nulliparious pregnancies. To evaluate these pregnancies, we train separate models on nulliparous patients. For training, we replace case report form features relating to delivery history with empty values. Results from training on nulliparous patients only are reported in Table 2.

**Table 2:** Comparisons of models trained only on nulliparous pregnancies. AUROC, AUPRC, and specificity at 90%17sensitivity highlighted for models trained on nulliparous data, grouped by which sources of data were included. Averages for AUROC and AUPRC are obtained by pooling classifier results together, and 95% confidence intervals are obtained by averaging all folds.

| Model | Ablation | AUROC |  | AUPRC |  | Spec. at 90% Sens. |
| --- | --- | --- | --- | --- | --- | --- |
|  |  | Pooled | 95% CI | Pooled | 95% CI |  |
| Gaussian Naive Bayes | Actigraphy Features | **0.719** | **0.723 (0.657 - 0.789)** | **0.183** | **0.291 (0.247 - 0.335)** | 0.448 (0.249 - 0.647) |
| Linear SVC | Actigraphy Features | 0.381 | 0.495 (0.326 - 0.664) | 0.085 | 0.210 (0.156 - 0.264) | 0.233 (-0.006 - 0.472) |
| Logistic Regression | Actigraphy Features | 0.551 | 0.727 (0.663 - 0.791) | 0.122 | 0.280 (0.245 - 0.315) | **0.460 (0.266 - 0.654)** |
| Nonlinear SVC | Actigraphy Features | 0.531 | 0.592 (0.493 - 0.691) | 0.108 | 0.245 (0.191 - 0.299) | 0.218 (0.089 - 0.348) |
| XGBoost | Actigraphy Features | 0.569 | 0.625 (0.568 - 0.682) | 0.116 | 0.315 (0.246 - 0.384) | 0.148 (-0.026 - 0.322) |
| Gaussian Naive Bayes | All Features | **0.590** | 0.602 (0.505 - 0.699) | **0.408** | **0.399 (0.319 - 0.479)** | 0.128 (-0.027 - 0.283) |
| Linear SVC | All Features | 0.377 | 0.456 (0.312 - 0.599) | 0.084 | 0.215 (0.169 - 0.261) | 0.158 (0.028 - 0.289) |
| Logistic Regression | All Features | 0.573 | **0.736 (0.665 - 0.806)** | 0.128 | 0.284 (0.248 - 0.320) | **0.426 (0.243 - 0.609)** |
| Nonlinear SVC | All Features | 0.472 | 0.575 (0.514 - 0.637) | 0.098 | 0.221 (0.181 - 0.261) | 0.246 (0.065 - 0.427) |
| XGBoost | All Features | 0.564 | 0.627 (0.535 - 0.719) | 0.125 | 0.331 (0.252 - 0.409) | 0.148 (-0.025 - 0.321) |
| Gaussian Naive Bayes | CRF Features | **0.492** | 0.502 (0.439 - 0.565) | **0.429** | **0.438 (0.388 - 0.487)** | 0.079 (0.012 - 0.147) |
| Linear SVC | CRF Features | 0.374 | 0.493 (0.353 - 0.633) | 0.085 | 0.263 (0.200 - 0.325) | 0.116 (0.009 - 0.224) |
| Logistic Regression | CRF Features | 0.444 | **0.714 (0.624 - 0.803)** | 0.113 | 0.325 (0.259 - 0.391) | **0.295 (0.066 - 0.524)** |
| Nonlinear SVC | CRF Features | 0.387 | 0.528 (0.407 - 0.649) | 0.094 | 0.225 (0.186 - 0.265) | 0.230 (0.023 - 0.437) |
| XGBoost | CRF Features | 0.528 | 0.609 (0.554 - 0.663) | 0.135 | 0.301 (0.230 - 0.372) | 0.047 (-0.061 - 0.155) |

As seen in Table 2, we find that the performance of actigraphy features is distinctive when we use Gaussian Näıve Bayes as the classifier. For all remaining model types, the performance is comparable both in contrast or together with case report form data, differing by relatively small amount for both AUROC and specificity at 90% sensitivity. This indicates that actigraphy data may provide performance comparable to or better than what can be assessed in a clinical survey specifically with regards to nulliparous patients.

In addition, the actigraphy features become a larger component of the most impactful features, as seen in Figure 8, although part of this can be attributed to the reduced dimensionality. Of the case report form features included, those relating to socioeconomic status appear to be the most impactful. When examining the actigraphy features only, as seen in Figure 9, we find that features evaluating variability are still among the most impactful features, whether they are averages of the day-to-day variability features or variances of the daily features.

**Figure 8:**
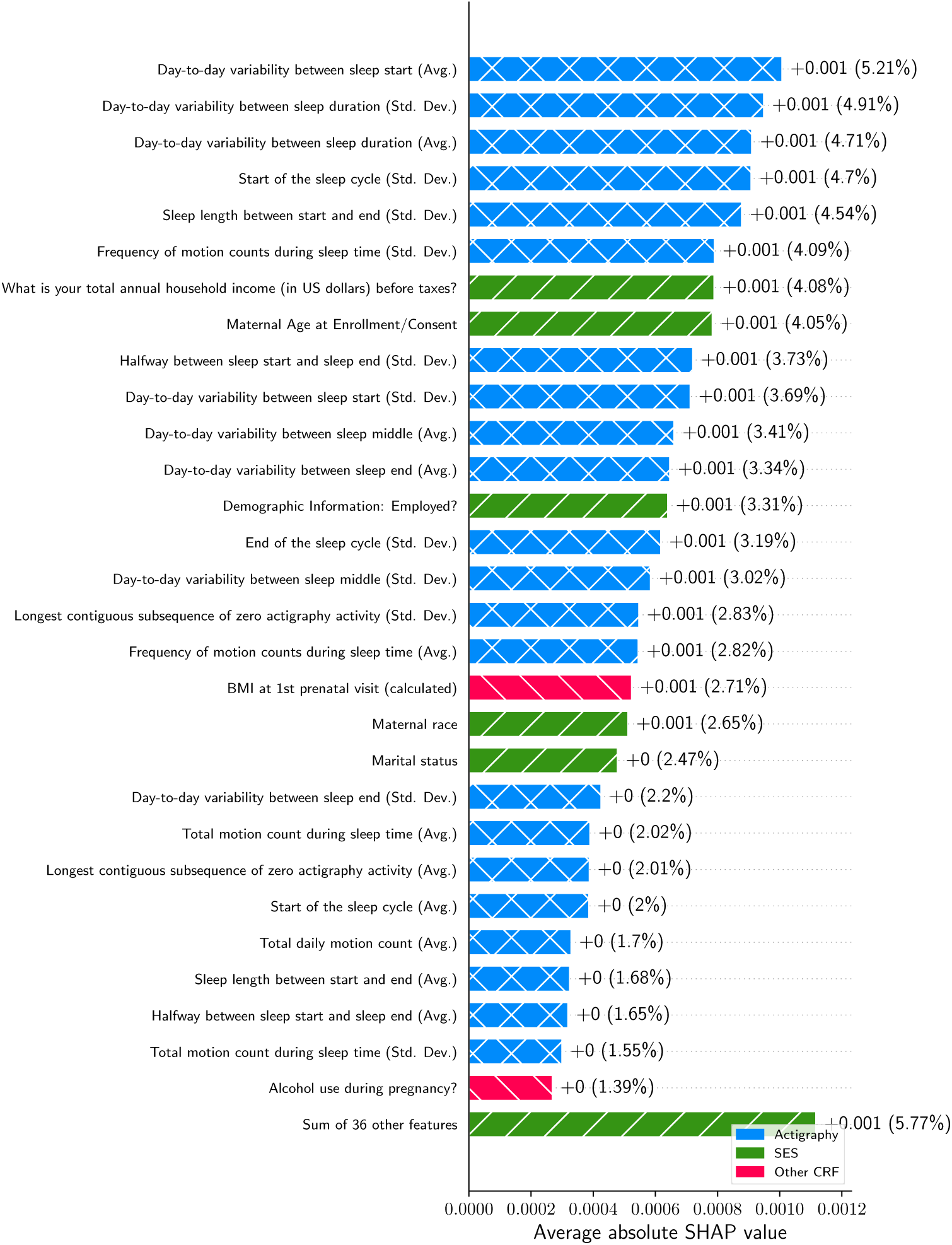
SHAP analysis of logistic regression with all features for nulliparous patients. Features indicative of socioeconomic status are highlighted green, and other patient history variables are highlighted red.

**Figure 9:**
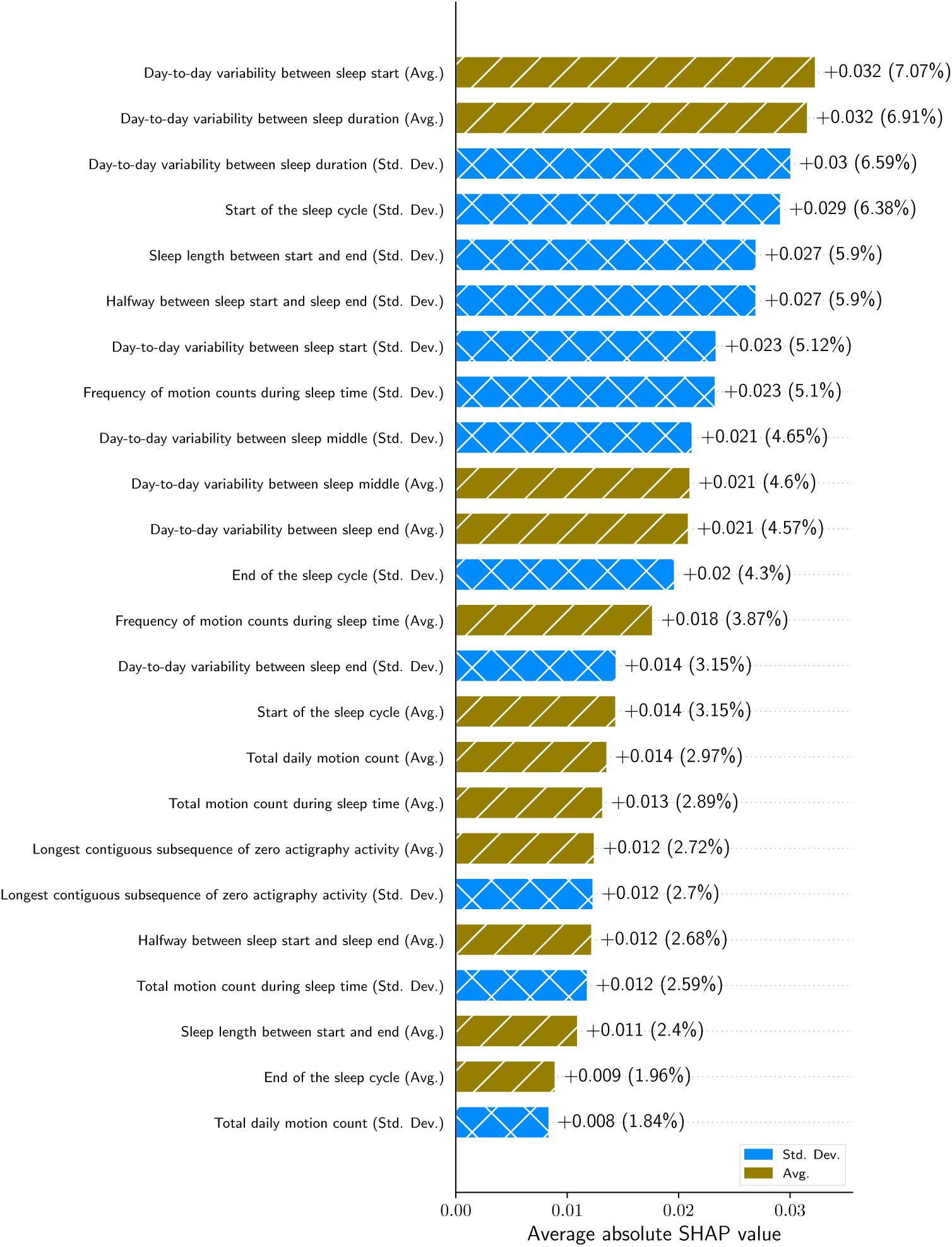
SHAP analysis of Gaussian NB with actigraphy features only. Features aggregating average patient behavior are highlighted in gold, and features aggregating the standard deviation of patient behaviors are highlighted in blue.

## 3 Discussion

Overall, we find that actigraphy data compiled into simple measures of sleep can aid in the prediction of PTB for both multiparous and nulliparous patients, and that simpler ML architectures appear to perform better at this. We also observe that features relating to sleep variability tend to rank highest among actigraphy features.

For all ablations tested, we find that Gaussian Näıve Bayes (Gaussian NB) has the highest average pooled AUROC. This is remarkable since it is architecturally simpler than other models, and suggests that the underlying features exhibit some independence from each other. This independence argument is furthered by the lower performance from our XGBoost models, as they learn decision trees where learned relationships may have dependencies. We do note that the small sample size and reduced dimensionality may enable this difference.

We also find that for the actigraphy-only models, there is a noticeable split in the explanainability between aggregating variability and averages of actigraphy features. Among the highest performing features, we find that those capturing variability in sleep patterns—either at the day-to-day or whole-sample level—were the most explainative features. Conversely, features examining a patient’s average behavior generally ranked lower, which suggests that consistent sleep patterns are more important than any specific sleep metric.

For nulliparous patients, we find that the overall performance of the actigraphy data is more comparable in performance to models trained on the case report form data only. When compared to whole-cohort models, the performance is similar for the actigraphy-based models, while the performance of the models trained on the case report form data drops noticeably. In addition to past PTB being a strong predictor of future PTB, this may suggest that monitoring sleep patterns is more necessary for nulliparous patients.

One limitation of this approach is that we do not evaluate categorical features as one-hot values, as the sample size would not be able to counterbalance the large number of features generated by one-hot categorical features. As a result, it is more difficult to interpret the impact of some categorical variables that do not actually have an ordinality to them (e.g. race, marital status). Similarly, we discard non-numerical features from the case report form features, as incorporating them with vision/language models would significantly increase the overall dimensionality; future models may incorporate these for improved performance. Moreover, we only evaluated mean imputation, further studies may evaluate the performance of other methods of imputation.

Sample size, particularly with regards to the nulliparous pregnancies, is another limiting issue, as it makes noise more prominent when training and evaluating these models. To mitigate this issue, we employed multiple random shufflings of the data for training and evaluation. Sample size is a limitation not only in cohort size, but also the amount of actigraphy data, as the duration and frequency at which study participants wore their actigraphy watches was not consistent. Further studies should evaluate larger cohorts of patients to ensure accurate performance measurements, as well as cohorts from other locations to validate the performance with respect to different demographics. Moreover, longer and more consistent usage of actigraphy watches may also reveal more reliable patterns of motion behavior that predict PTB.

In addition, future work with actigraphy data could incorporate luminosity sensor data, as it may provide additional signals and corroborate signals captured by an actigraphy sensor.

Another area of future work are with models trained with self-supervised learning (SSL), which learn relationships between input features before being fine-tuned for a downstream task. SSL models are particularly effective as these learned relationships between features generalize well in supervised tasks [19].

## 4 Methods

### 4.1 Study Characteristics

This study was completed as a part of of the March of Dimes Prematurity Research Center at Washington University in St. Louis/BJC HealthCare [12], which was approved by the Washington University IRB (reference #201612070) in accordance FDA Good Clinical Practices and the Declaration of Helsinki. Written informed consent was obtained from participants for the usage of their clinical, biospecimen, imaging, and questionnaire data. Patients were recruited at the Washington University Medical Campus if they had a singleton pregnancy with an estimated GA under 20 weeks, planned to deliver at Barnes-Jewish Hospital, and were age 18 or older.

Trained obstetric research staff used a series of case report forms to collect baseline maternal demographics, medical history, antepartum data and obstetric outcomes as previously described in [12]. Patient data were collected at scheduled study visits during each trimester and at delivery, where biological samples, imaging, actigraphy, and responses to standardized surveys were obtained from each patient.

Survey data included questions from eleven different validated surveys and standalone questions covering stress, schedule, sleep quality, physical activity, postnatal depression, diet, demographics, and overall lifestyle. We derive the label of PTB from the reported estimated date of confinement (EDC), labeling births that occur 3 full weeks before the listed EDC as PTB. EDC was derived from the patient’s last menstrual period or first ultrasound [20].

### 4.2 Actigraphy Feature Design

Actigraphy measurements were collected over a two-week period in each trimester (first trimester: 0 to 13 weeks and 6 days, second trimester: 14 weeks to 27 weeks and 6 days, third trimester: *≥* 28 weeks) using the CamNtech MotionWatch 8. Measurements were collected at a minute-frequency over the duration a study participant wore their actigraphy watch. Patients were reminded through calls, emails, and texts to return their actigraphy watches after the capture period either at the next study visit or through a courier [12]. Patients who did not have actigraphy data in either their first or second trimester were filtered from the results for this analysis.

These features are very high-resolution, and to ensure the data is tractable for shallow ML model training, we engineer these raw time-series signals into aggregate features over day-level windows. On top of the day-level measurements, we also measure the absolute change between days where data is present. Table 3 contains a summary of these engineered features.

**Table 3:**
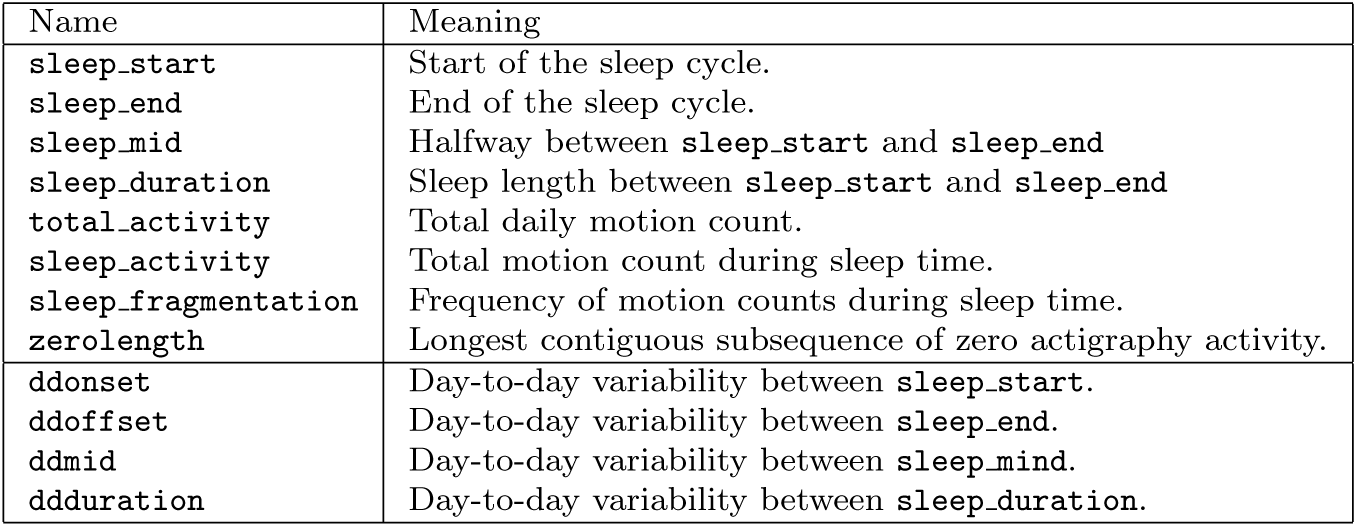
Summary of the engineered actigraphy features and their meanings.

| Name | Meaning |
| --- | --- |
| `sleep_start` | Start of the sleep cycle. |
| `sleep_end` | End of the sleep cycle. |
| `sleep_mid` | Halfway between `sleep_start` and `sleep_end` |
| `sleep_duration` | Sleep length between `sleep_start` and `sleep_end` |
| `total_activity` | Total daily motion count. |
| `sleep_activity` | Total motion count during sleep time. |
| `sleep_fragmentation` | Frequency of motion counts during sleep time. |
| `zerolength` | Longest contiguous subsequence of zero actigraphy activity. |
| `ddonset` | Day-to-day variability between `sleep_start`. |
| `ddoffset` | Day-to-day variability between `sleep_end`. |
| `ddmid` | Day-to-day variability between `sleep_mid`. |
| `ddduration` | Day-to-day variability between `sleep_duration`. |

To generate these features, all actigraphy data is separated into days centered around midnight, from which we then attempt to estimate the sleep cycle that occurred for each given day. Table 3 gives a summary of the calculated features that were used in the dataset for the ML models.

### 4.3 Model Design

For each study participant, we aggregate the day-level actigraphy features down to their mean and standard deviation across the entire duration of the pregnancy. When evaluating the window of GAs below a full-term pregnancy, we drop all actigraphy data with a GA below a set range (*e.g.* if we set the upper limit at 140 days, all data before 140 days are dropped, and the remainder is aggregated).

For the survey data, we select features with both domain knowledge and automatic techniques. We first select a predefined set of features based on pre-determined clinical knowledge, and sum values of questions regarding individual births together. After these features, we select an additional 10 features with the minimal-redundancy-maximal-relevance (mRMR) algorithm with semantic textual similarity (STS) scores generated with PubMedBERT [21] fine-tuned on several clinical and general datasets, as described in [22]. Features not represented numerically are dropped. The full list of features that we used can be found in Section 2 of the Supplementary Materials.

After this, we concatenate both sources of data, scale all numerical features to its normal distribution, and encode all categorical features as ordinal values. Missing values are imputed with the mean of the feature. The data is randomly split across a 80%/20% train/test split. For the whole cohort, 532 and 133 patients appear in in each split. For the nulliparous cohort, this becomes 238 in the train set and 59 in the test set. A summary of the training pipeline can be seen in Figure 10.

**Figure 10:**
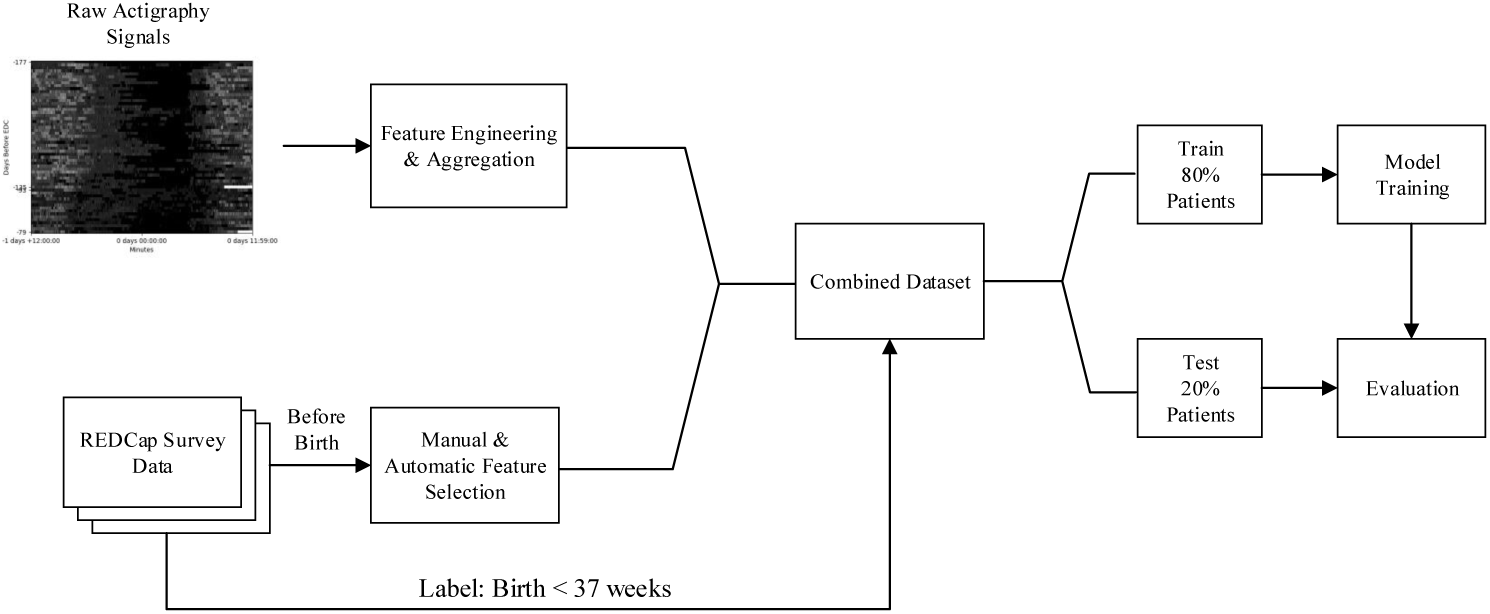
Diagram of data and model pipeline used in this study. The dataset is combined from two primary sources of data, raw actigraphy signals and Research Electronic Data Capture (REDCap) survey data. The raw actigraphy signals are preprocessed as engineered features and aggregated by day-level results. Manual and automatic feature selection is applied to the REDCap survey data. The combined dataset is split into 80% of training data for the patients, and 20% for the evaluation of the trained models.

We train our models with several standard ML models, including XGBoost [23], linear support vector machine (SVM), kernelized/non-linear SVM [24], logistic regression, and Gaussian NB [25]. We evaluate the results across 10 random initializations for each model in Section 2, and report the average AUROC and AUPRC through pooling [26], as well as the 95% confidence interval over all initializations. SHAP values are averaged across all random initializations.

To find the best hyperparameters for each of the tested models, we use 5-fold flat cross-validation (CV) across the training set. For XGBoost, the hyperparameter space ranges from 1 to 3 estimators, 1 to 3 maximum depth, a learning rate of 0.1, and a fitting objective of AUROC. For linear SVM, we test regularization parameters ranging logarithmically from 0.001 to 10, with 1000 iterations of training. For non-linear SVM, we evaluate polynomial and radial basis function kernels on top of the linear SVM parameters. For logistic regression, we evaluate regularization parameters from 0.001 to 10 with a *L*_2_ penalty, and 1000 maximum iterations of training. For Gaussian NB, we use 10*^−^*^9^ as a fixed smoothing parameter.

## Data Availability

The data used in these findings can be obtained from the authors by request with permission from Washington University in St. Louis.

## Code Availability

We make the code used in this study available at https://github.com/bcwarner/mod-actigraphy-clf.

## Acknowledgements

We thank the participants in the cohort study for their key contributions in advancing preterm birth science, as well as the staff who facilitated the enrollment of participants and collection of data that made this study possible.

This work was supported financially by a research grant from the March of Dimes Foundation, and was supported institutionally by St. Louis Children’s Hospital, Barnes-Jewish Hospital, Washington University School of Medicine in St. Louis, and the Washington University in St. Louis James McKelvey School of Engineering.

## Author Contributions

B.C.W. wrote the main manuscript text and contributed the code for the training, evaluation and interpretation of the models. P.Z. contributed the code for the features. E.D.H., S.K.E. A.I.F., and C.L. provided subject matter expertise. All authors were responsible for the interpretation of the models.

## References

1. Cao, G., Liu, J. & Liu, M. Global, Regional, and National Incidence and Mortality of Neonatal Preterm Birth, 1990-2019. JAMA Pediatrics 176, 787–796 (2022). URL https://www.ncbi.nlm.nih.gov/pmc/articles/PMC9157382/.

2. Chawla, D. & Agarwal, R. Preterm births and deaths: from counting to classification. The Lancet Global Health 10, e1537–e1538 (2022). URL https://linkinghub.elsevier.com/retrieve/pii/S2214109X22004223.

3. Sutcliffe, S. et al. Risk of pre-term birth as a function of sleep quality and obesity: prospective analysis in a large Prematurity Research Cohort. Sleep Advances 4, zpad043 (2023). URL https://academic.oup.com/sleepadvances/article/doi/10.1093/sleepadvances/zpad043/7337357.

4. Wang, L. & Jin, F. Association between maternal sleep duration and quality, and the risk of preterm birth: a systematic review and meta-analysis of observational studies. BMC Pregnancy and Childbirth 20, 125 (2020). URL https://bmcpregnancychildbirth.biomedcentral.com/articles/10.1186/s12884-020-2814-5.

5. Hoyniak, C. P. et al. The Association Between Maternal Sleep and Circadian Rhythms During Pregnancy and Infant Sleep and Socioemotional Outcomes (2024). URL https://www.researchsquare.com/article/rs-3937599/v1.

6. Hoyniak, C. P. et al. Sleep and circadian rhythms during pregnancy, social disadvantage, and alterations in brain development in neonates. Developmental Science 27, e13456 (2024). URL https://onlinelibrary.wiley.com/doi/abs/10.1111/desc.13456. eprint: https://onlinelibrary.wiley.com/doi/pdf/10.1111/desc.13456.

7. Li, R. et al. Sleep disturbances during pregnancy are associated with cesarean delivery and preterm birth. The Journal of Maternal-Fetal & Neonatal Medicine 30, 733–738 (2017). URL https://doi.org/10.1080/14767058.2016.1183637. Publisher: Taylor & Francis eprint: 10.1080/14767058.2016.1183637.

8. Cespedes, E. M. et al. Comparison of Self-Reported Sleep Duration With Actigraphy: Results From the Hispanic Community Health Study/Study of Latinos Sueno Ancillary Study. American Journal of Epidemiology 183, 561–573 (2016). URL https://academic.oup.com/aje/article-lookup/doi/10.1093/aje/kwv251.

9. Nauha, L. et al. Comparison and agreement between device-estimated and self-reported sleep periods in adults. Annals of Medicine 55, 2191001 (2023). URL https://doi.org/10.1080/07853890.2023.2191001. Publisher: Taylor & Francis eprint: 10.1080/07853890.2023.2191001.

10. Cos, H. et al. Predicting outcomes in patients undergoing pancreatectomy using wearable technology and machine learning: prospective cohort study. Journal of medical Internet research 23, e23595 (2021). URL https://www.jmir.org/2021/3/e23595/. Publisher: JMIR Publications Toronto, Canada.

11. Dai, R. et al. Multi-Task Learning for Randomized Controlled Trials: A Case Study on Predicting Depression with Wearable Data. Proceedings of the ACM on Interactive, Mobile, Wearable and Ubiquitous Technologies 6, 1–23 (2022). URL https://dl.acm.org/doi/10.1145/3534591.

12. Stout, M. J. et al. A multidisciplinary Prematurity Research Cohort Study. PLOS ONE 17, e0272155 (2022). URL https://dx.plos.org/10.1371/journal.pone.0272155.

13. Ravindra, N. G. et al. Deep representation learning identifies associations between physical activity and sleep patterns during pregnancy and prematurity. npj Digital Medicine 6, 171 (2023). URL https://www.nature.com/articles/s41746-023-00911-x.

14. Lundberg, S. M. & Lee, S.-I. A unified approach to interpreting model predictions. Advances in neural information processing systems 30 (2017).

15. Varner, M. W. & Esplin, M. S. Current understanding of genetic factors in preterm birth. BJOG: An International Journal of Obstetrics & Gynaecology 112, 28–31 (2005). URL https://onlinelibrary.wiley.com/doi/abs/10.1111/j.1471-0528.2005.00581.x. eprint: https://onlinelibrary.wiley.com/doi/pdf/10.1111/j.1471-0528.2005.00581.x.

16. Hsieh, T.-T. et al. The Impact of Interpregnancy Interval and Previous Preterm Birth on the Subsequent Risk of Preterm Birth. Journal of the Society for Gynecologic Investigation 12, 202–207 (2005). URL http://journals.sagepub.com/doi/10.1016/j.jsgi.2004.12.004.

17. Culhane, J. F. & Goldenberg, R. L. Racial Disparities in Preterm Birth. Seminars in Perinatology 35, 234–239 (2011). URL https://linkinghub.elsevier.com/retrieve/pii/S0146000511000498.

18. Goldenberg, R. L., Culhane, J. F., Iams, J. D. & Romero, R. Epidemiology and causes of preterm birth 371 (2008).

19. Erhan, D., Courville, A., Bengio, Y. & Vincent, P. Why Does Unsupervised Pre-training Help Deep Learning? Proceedings of the Thirteenth International Conference on Artificial Intelligence and Statistics 201–208 (2010). URL https://proceedings.mlr.press/v9/erhan10a.html. ISSN: 1938-7228.

20. Clinical Management Guidelines for Obstetrician–Gynecologists. Number 49, December 2003: (Replaces Technical Bulletin Number 218, December 1995). Obstetrics & Gynecology 102, 1445–1454 (2003). URL http://journals.lww.com/00006250-200312000-00048.

21. Gu, Y. et al. Domain-specific language model pretraining for biomedical natural language processing. ACM Transactions on Computing for Healthcare (HEALTH) 3, 1–23 (2021). Publisher: ACM New York, NY.

22. Warner, B. C., Xu, Z., Haroutounian, S., Kannampallil, T. & Lu, C. Utilizing Semantic Textual Similarity for Clinical Survey Data Feature Selection (2023). URL http://arxiv.org/abs/2308.09892. ArXiv:2308.09892 [cs].

23. Chen, T. & Guestrin, C. XGBoost: A Scalable Tree Boosting System. Proceedings of the 22nd ACM SIGKDD International Conference on Knowledge Discovery and Data Mining 785–794 (2016). URL https://dl.acm.org/doi/10.1145/2939672.2939785.

24. Cortes, C. & Vapnik, V. Support-vector networks. Machine Learning 20, 273–297 (1995). URL http://link.springer.com/10.1007/BF00994018.

25. Chan, T. F., Golub, G. H. & LeVeque, R. J. in *Updating Formulae and a Pairwise Algorithm for Computing Sample Variances* (eds Caussinus, H., Ettinger, P. & Tomassone, R.) COMPSTAT 1982 5th Symposium held at Toulouse 1982 30–41 (Physica-Verlag HD, Heidelberg, 1982). URL http://link.springer.com/10.1007/978-3-642-51461-6 3.

26. Jack, H. & Niall, A. On Averaging ROC Curves. Transactions on Machine Learning Research (2023). URL https://openreview.net/forum?id=FByH3qL87G.

